# Estimates of COVID-19 case-fatality risk from individual-level data

**DOI:** 10.1101/2020.04.16.20067751

**Authors:** Simona Bignami-Van Assche, Daniela Ghio, Ari Van Assche

## Abstract

When calculated from aggregate data on confirmed cases and deaths, the case-fatality risk (CFR) is a simple ratio between the former and the latter, which is prone to numerous biases. With individual-level data, the CFR can be estimated as a true measure of risk as the proportion of incidence for the disease. We present the first estimates of the CFR for COVID-19 by age and sex based on event history modelling of the risk of dying among confirmed positive individuals in the Canadian province of Ontario, which maintains one of the few individual-level datasets on COVID-19 in the world.

## INTRODUCTION

The severity profile of a novel pathogen is one of the most critical issues as it begins to spread, when assessing disease course and outcome is crucial for planning health interventions (*1*). For this reason, an important epidemiological indicator to monitor during the current outbreak of COVID-19 is the case-fatality risk (CFR), the proportion of confirmed cases who result in fatalities. When calculated from aggregate data on confirmed cases and deaths, the CFR is a simple ratio between the former and the latter, which is prone to numerous biases (*1*–*5*). With individual-level data, the CFR can be estimated as a true measure of risk as the proportion of incidence for the disease.

There are only two accessible individual-level datasets on COVID-19 confirmed positive cases that include their clinical outcome (death or recovery), which are maintained by the Canadian provinces of Ontario and Alberta^1^. Ontario is currently the second province in Canada for number of COVID-19 infections and fatalities. We present the first estimates of the CFR for COVID-19 by age and sex based on event history modelling of the risk of dying among confirmed positive individuals in Ontario.^2^

## METHODS

Between January 1 and April 15, 2020, Ontario recorded 8,447 confirmed positive cases of COVID-19, of which 3,902 had recovered and 385 had died. The individual-level dataset we analyse contains basic information on age, gender, clinical outcome (death or recovery) and name of the reporting health facility for these cases. Individuals’ age is recorded at the time of testing in ten-years groups, with information missing for 5 cases (of which 2 are recoveries and 3 have an unresolved outcome). Information on gender is missing for 46 cases (of which 5 are fatalities, 12 are recoveries, and 29 have an unresolved outcome). These observations were excluded, so that the analysis is based on 8,394 cases, of whom 56.2% (4,716) are females.

Following the guidelines of the World Health Organization, in Canada only individuals symptomatic with fever, cough and/or difficulty breathing are tested for COVID-19. In Ontario, case definition refers to these symptomatic individuals who tested positive for the virus and also to their close contacts even if they were not tested for the disease. In our dataset, this latter category included 1,429 (17%) of cases.

We estimate the CFR through event history modelling in order to take into account censoring that arises because, at the time of observation, the outcome is unknown for a nonnegligible portion of infected individuals. In this framework, the CFR coincides with the cumulative incidence (CI) of mortality, with recovery as a competing risk. The CI is estimated after fitting the competing risk regression model, controlling for gender, with *stcrreg* in Stata/SE (version 12.0, StataCorp, LLC).

## RESULTS

Of 8,394 confirmed positive cases considered, 191 (2.3%) were under 20 years age; 946 (11.3%) age 20-29; 1,045 (12.5%) age 30-39; 1,241 (14.8%) age 40-49; 1,545 (18.4%) age 50-59; 1,188 (14.2%) age 60-69; 794 (9.5%) age 70-79; 873 (10.4%) age 80-89; and 571 (6.8%) age 90 or more.

The CFR (cumulative incidence of mortality vs recovery) for COVID-19 confirmed positive cases in Ontario is presented in Table 1. It can be seen that it increases exponentially with age, reaching 18.7% for those aged 90+. By comparing the estimated cumulative incidence of death for COVID-19 with the crude case-fatality risk calculated from aggregate data on the total number of cases and deaths, we can appreciate to what extent the latter over-estimates the risk of COVID-19-related mortality in each age group. This is especially the case in the 70-79 and 80-89 age groups, when the difference between the two estimates is 6.2% and 5.0%, respectively.

**Table 1.** Cumulative incidence of mortality (case-fatality risk) from individual-level data, with recovery as competing risk, among COVID-19 confirmed positive cases in Ontario as of April 15, by age, and case-fatality ratio from aggregate data on cases and deaths Cumulative incidence is estimated controlling for patients’ gender.

| Age group | No. at risk | Cumulative incidence, % | Case-fatality ratio, % |
| --- | --- | --- | --- |
| <20 | 8394 | 0 | 0 |
| 20-29 | 8203 | 0 | 0 |
| 30-39 | 7257 | 0 | 0.1 |
| 40-49 | 6212 | 0.2 | 0.6 |
| 50-59 | 4971 | 0.6 | 1.0 |
| 60-69 | 3426 | 1.4 | 2.4 |
| 70-79 | 2238 | 4.5 | 10.7 |
| 80-89 | 1444 | 10.5 | 15.5 |
| 90+ | 571 | 18.7 | 18.9 |
| Total | --- | 3.1 | 4.5 |

At any age, female positive cases have a risk of mortality that is 29 percent lower than male cases (SHR=.706; CI=.581, .858; *p*=.000). The comparison of the cumulative incidence of COVID-19-related mortality for males and females is presented in Figure 1.

**Figure 1.**
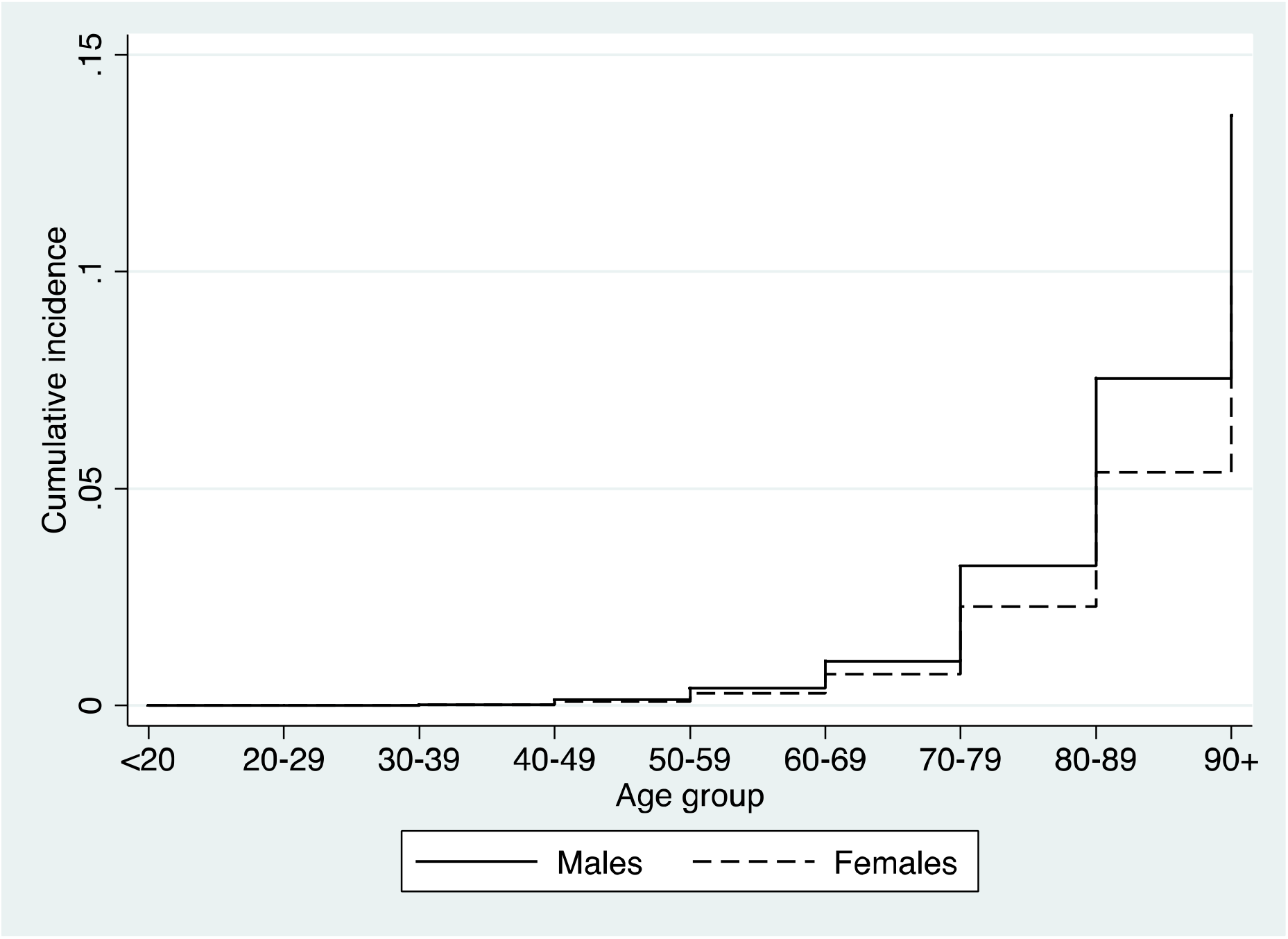
Cumulative incidence of mortality (case-fatality risk) from individual-level data, with recovery as competing risk, among COVID-19 confirmed positive cases in Ontario as of April 15, by sex

## CONCLUSIONS

Our results show that the CFR for COVID-19 can be more accurately estimated with individual-level data. This is important not only to understand the evolution of the pandemic, but also to evaluate the effectiveness of new treatments as they are introduced. Information on patients’ medical history would improve the analysis by discounting the effect of pre-existing conditions on the risk of mortality for COVID-19. Utilization of health care resources could be also assessed with information on hospitalizations and on the time elapsed between diagnosis and clinical outcome. As we and several scholars indicated (*6*-*7*), the lack of comparable data and indicators is hampering our efforts to win the war against COVID-19. National and global coordination in this area is urgently needed.

## Data Availability

Data are available through the website of the Government of Ontario.

https://data.ontario.ca/dataset?keywords_en=COVID-19.

## Footnotes

1 The dataset for Ontario is available at: https://data.ontario.ca/dataset?keywords_en=COVID-19. The dataset for Alberta at: https://covid19stats.alberta.ca.

2 The number of COVID-19-related deaths recorded in Alberta (48 as of April 15) is too small for this type of analysis.

